# Innovating Urban Mosquito Control: Introducing Human-Controlled Breeding Sites as a Component of Integrated Mosquito Management (IMM)

**DOI:** 10.1101/2024.05.14.24307332

**Authors:** Guillermo Antonio Baigorria, Consuelo Cecilia Romero

## Abstract

**Background:** Urbanization and climate change exacerbate mosquito-borne disease transmission by expanding mosquito habitats in densely populated areas. Traditional mosquito control methods, such as eliminating breeding sites or using insecticides, face challenges in urban settings due to practicality and resistance issues. This study introduces Human-Controlled Breeding Sites (HCBS), a novel, low-cost, and environmentally friendly mosquito control method tailored for urban and suburban households. HCBS provides controlled oviposition sites to attract gravid female mosquitoes, disrupting their reproductive cycle without chemical deterrents.

**Methods:** The HCBS method involves deploying devices designed to mimic natural breeding sites, encouraging mosquitoes to lay eggs in a controlled environment. After 10 days, eggs, larvae, and pupae are collected and destroyed. The HCBS device, fabricated using 3D-printed ABS material in five colors (white, black, red, green, blue), was tested in a semi-controlled laboratory in La Molina, Peru. The experiment evaluated oviposition preferences of *Aedes aegypti* by counting larvae and pupae across five 10-day cycles, with device colors randomized to minimize bias.

**Results:** HCBS devices successfully attracted gravid *Aedes aegypti* for oviposition, with green devices showing the highest initial preference (2-17 larvae/pupae per cycle), though larvae were later observed in all colors, indicating color is not essential. The method achieved 100% effectiveness in disrupting the mosquito life cycle by eliminating eggs, larvae, and pupae within the 10-day timeframe. The low-cost design and use of household materials enhance its scalability and community engagement potential.

**Conclusions:** HCBS offers a sustainable alternative to conventional mosquito control by targeting early life stages, reducing populations with minimal environmental impact. Its adaptability to household settings and cost-effectiveness make it suitable for urban and suburban areas. Further research is needed to optimize device design, assess long-term efficacy, and integrate HCBS into disaster risk management systems for broader public health applications, particularly in regions like Peru with documented insecticide resistance.

## **1.** INTRODUCTION

Climate change has significantly impacted mosquito population dynamics, leading to the expansion of mosquito habitats into new regions due to rising temperatures, altered precipitation, and humidity changes [1]. This shift increases the risk of mosquito-borne diseases in previously unaffected areas [2]. Urbanization further exacerbates this risk by creating environments conducive to mosquito proliferation, particularly in densely populated areas with abundant breeding sites [3,4]. The convergence of high human density and expanding mosquito populations in urban settings amplifies the transmission of diseases like Dengue, Malaria, West Nile virus, yellow fever, and Zika [5].

Traditional mosquito control methods, designed for rural areas, face challenges in urban environments due to the impracticality of widespread insecticide use and the limited effectiveness of biological controls [6]. Urban landscapes also complicate source reduction and habitat modification strategies. Addressing the growing threat of mosquito-borne diseases in urban areas requires innovative, context-specific control methods. Integrated approaches that combine novel vector control technologies with community engagement and public health interventions offer promising solutions [7]. These may include new larvicides, spatial repellents, and community-based surveillance systems.

The objectives of this study are: (1) to introduce a novel method for mosquito control specifically designed for urban and suburban areas, addressing the unique challenges these environments present; (2) to present the concept of Human-Controlled Breeding Sites (HCBS) as an innovative approach to mosquito population reduction by providing a controlled environment for mosquito females to oviposit their eggs within households; (3) to describe the design and functionality of the HCBS device, emphasizing its potential as a supplementary component of Integrated Mosquito Management (IMM) strategies; and (4) to investigate the oviposition preferences of mosquito females by testing multiple colors within the HCBS devices, aiming to identify the most attractive color for egg-laying and optimize the effectiveness of the control method.

This paper is structured into three main components. Section 3.1 details the meticulous development of the HCBS method, laying the foundational groundwork for substantiating the subsequent exploration and validation of the HCBS methodology. Section 3.2 delves into the intricate development of the HCBS device, an essential component for implementing the HCBS approach effectively. Finally, Section 3.3 is dedicated to describing the experiment conducted to validate the HCBS methodology, offering insights into its practical application and efficacy in mosquito population control. Together, these integrated components provide a comprehensive understanding of the HCBS approach, effectively bridging the gap between methodological development, device design, and practical application.

## 2. LITERATURE REVIEW

Mosquito-borne diseases pose significant threats to public health, particularly in tropical and subtropical regions worldwide. Among the most notorious mosquito-borne illnesses are Dengue fever, Malaria, Yellow Fever, West Nile virus and Zika, which collectively cause millions of cases and thousands of deaths each year [8]. Controlling the populations of disease-carrying mosquitoes is crucial for reducing the transmission and burden of these diseases. In this literature review, we explore the biology and behavior of mosquitoes, existing control methods, factors influencing mosquito attraction, and the relationship between mosquito species and disease transmission.

### 2.1 Mosquito Biology and Behavior

Mosquitoes, belonging to the Culicidae family, are small, flying insects characterized by their slender bodies, long legs, and elongated mouthparts adapted for piercing and sucking blood^9^. The life cycle of mosquitoes consists of four stages: egg, larva, pupa, and adult.

Male and female mosquitoes exhibit significant differences in their lifespans. Male mosquitoes typically live for up to 10 days [9,10], while females can survive for a few weeks [11]. Some sources suggest that males live for 1-2 weeks [12] and females typically live anywhere from two to four weeks [13].

The primary reason for this difference in lifespan is the distinct roles each gender plays within the species. Males primarily live to mate and do not play a major role in the reproductive cycle after mating [12]. On the other hand, female mosquitoes require extended periods of life to lay eggs [9].

Moreover, the feeding habits of male and female mosquitoes also contribute to their lifespan differences. Male mosquitoes feed only on plant nectar and sugary liquids, such as fruit juices and honeydew [9]. In contrast, female mosquitoes feed on blood in addition to plant nectar, which provides the necessary nutrients for egg production [11].

Therefore, the differences in lifespan between male and female mosquitoes can be attributed to their distinct roles in reproduction and their different feeding habits [9–11,13,14], although a new study suggest the lifespan difference can be less [10].

Environmental factors significantly influence mosquito biology and behavior, with temperature, humidity, and the availability of breeding sites playing crucial roles in shaping mosquito population dynamics and distribution [15,16]. Human-related factors, such as urbanization [17] and changes in land use, can also have a substantial impact on mosquito habitats and abundance [18,19]. Additionally, characteristics like water type [20], larvae density [21–24], and even pheromone emissions by conspecific larvae [20,25] influence oviposition behavior. However, a common thread among these factors is the necessity of water in all oviposition sites.

### 2.2 Mosquito Control Methods

The World Health Organization [26] advocates for integrated vector management (IVM) and suggests four major strategies: Chemical control involves the use of insecticides to kill mosquito larvae and adults. These variously comprise fogging, residual spraying, larviciding, and autodissemination. However, the emergence of insecticide resistance poses challenges to the effectiveness of chemical control efforts [27].

Biological control methods harness natural enemies of mosquitoes, such as predators and parasites, to reduce mosquito populations. Predatory fish, such as *Gambusia affinis* (mosquito fish), feed on mosquito larvae and are used in biological control programs [28]. Additionally, genetically modified mosquitoes are being developed as a novel approach to suppress mosquito populations [29].

Environmental management strategies focus on reducing mosquito breeding sites and modifying habitats to make them less suitable for mosquito development. Source reduction techniques involve eliminating or treating standing water where mosquitoes breed, while habitat modification aims to alter environmental conditions to discourage mosquito proliferation [30].

Personal barrier measures involve using barriers like window screens, mosquito nets, repellents, or protective clothing to prevent mosquito bites.

Integrated Mosquito Management (IMM) emphasizes the coordinated use of multiple control methods tailored to local conditions. Surveillance, larval control, adult control, and community engagement are key components of IMM programs aimed at reducing mosquito populations and disease transmission [31].

### 2.3 Mosquito Attraction and Repellents

Mosquitoes are attracted to their hosts by a combination of visual and chemical cues. Visual cues, such as color, play a role in mosquito attraction. Research suggests that mosquitoes are more attracted to dark colors, possibly due to their increased visibility against contrasting backgrounds [32].

Chemical cues, including carbon dioxide (CO2) and human body odors, also influence mosquito host-seeking behavior. Mosquitoes are highly sensitive to CO2, which is emitted by humans during respiration and serves as a potent attractant for female mosquitoes seeking blood meals [33]. Additionally, human skin odors, such as lactic acid and ammonia, can attract mosquitoes from a distance [34].

Various repellents are available to protect against mosquito bites. Synthetic chemicals, such as DEET (N,N-diethyl-meta-toluamide), are widely used and highly effective at repelling mosquitoes [35]. Plant-derived repellents, such as citronella and eucalyptus oil, offer alternative options for personal protection against mosquito bites [36].

### 2.4 Mosquito Species and Disease Transmission

Different mosquito species exhibit varying degrees of vector competence for transmitting diseases to humans. *Aedes aegypti* and *Aedes albopictus* are primary vectors of Dengue fever, Zika virus, West Nile virus and Chikungunya virus, with *Aedes aegypti* being particularly efficient in urban environments [37]. *Anopheles* mosquitoes are the main vectors of Malaria, transmitting Plasmodium parasites to humans through their blood meals [38].

The relationship between mosquito species and disease transmission is influenced by factors such as vector biology, host preferences, and pathogen development. Understanding the ecology and behavior of mosquito vectors is essential for implementing targeted control measures to reduce disease transmission [39].

### 2.5 Mosquito’s oviposition ethology

Mosquito oviposition behavior, influenced by various factors including color preferences and environmental cues, plays a crucial role in the transmission of diseases. Different mosquito species exhibit distinct preferences in color for oviposition sites, often correlating with the diseases they transmit. Understanding these ethological preferences is essential for effective vector control strategies and disease prevention efforts.

#### 2.5.1 Aedes aegypti

*Aedes aegypti* is a primary vector of several devastating diseases, including dengue fever, Zika virus, West Nile virus, and chikungunya. Research has shown that *Aedes aegypti* females exhibit a preference for dark colors such as black and brown for oviposition sites [40]. These dark-colored sites often contain stagnant water, such as discarded containers, which serve as breeding grounds for *Aedes* mosquitoes.

#### 2.5.2 Aedes albopictus

Another important vector species, *Aedes albopictus*, is associated with the transmission of diseases such as dengue fever, Zika virus, West Nile virus, and chikungunya. Unlike *Aedes aegypti*, *Aedes albopictus* displays a preference for lighter colors like white or light blue for oviposition [41]. These preferences may vary depending on environmental conditions and habitat characteristics, influencing the distribution of oviposition sites and disease transmission patterns.

#### 2.5.3 Culex pipiens

*Culex pipiens* is a widespread mosquito species known for transmitting West Nile virus, a potentially severe neurological disease transmitted to humans and other vertebrate hosts. Studies have indicated that *Culex pipiens* females prefer oviposition sites with high organic content, often found in dark-colored water bodies [42].

#### 2.5.4 Anopheles spp

*Anopheles* mosquitoes are vectors of malaria parasites, a life-threatening disease that affects millions of people worldwide. Different species within the *Anopheles* genus may exhibit varying oviposition preferences. For example, *Anopheles gambiae*, a major malaria vector in sub-Saharan Africa, shows a preference for dark-colored oviposition sites [43]. These sites, often found in natural water bodies like swamps and puddles, provide suitable breeding habitats for *Anopheles* mosquitoes and contribute to the transmission of malaria.

In addition to color preferences, environmental factors such as temperature, humidity, and vegetation also influence mosquito oviposition behavior and disease transmission dynamics. Mosquitoes prefer shaded areas with moderate temperatures and high humidity for oviposition, as these conditions promote larval survival and development [44]. Vegetation provides additional shelter and resources, creating ideal habitats for mosquito breeding and disease transmission ^9^.

#### 2.5.5 Oviposition radius

Gravid mosquito females exhibit a remarkable ability to search for suitable oviposition sites, often traveling significant distances and enduring varying environmental conditions to lay their eggs in standing water. The duration and distance traveled by gravid females largely depend on factors such as species, habitat availability, and physiological state. For instance, *Aedes* mosquitoes, vectors of diseases like Dengue and Zika, have been observed to travel up to several hundred meters in search of suitable breeding sites, with some individuals capable of flying distances of over a kilometer [45]. Similarly, *Anopheles* mosquitoes, responsible for transmitting malaria, are known to actively seek out water bodies for oviposition, often traveling several kilometers from their blood-feeding sites [9]. Gravid females may spend several days or even weeks in search of suitable oviposition sites, utilizing various sensory cues and environmental cues to guide their behavior [9]. This remarkable behavior underscores the importance of effective mosquito control strategies to target breeding sites and minimize the risk of disease transmission.

#### 2.5.6 Mosquito eggs viability

Mosquito eggs possess remarkable resilience, capable of surviving in a desiccated state for extended periods. Research indicates that mosquito eggs can remain viable for varying durations without water, depending on species and environmental conditions. For instance, Aedes mosquitoes, known vectors of diseases such as Dengue and Zika, have been reported to maintain viability for up to several months under favorable conditions [46]. Similarly, studies on *Anopheles* mosquitoes, responsible for transmitting malaria, suggest that their eggs can endure desiccation for several weeks, retaining the potential for hatching upon rehydration [45]. This adaptive trait enables mosquito eggs to persist through adverse environmental conditions, contributing to the resilience and persistence of mosquito populations.

Mosquito eggs are typically laid on the surface of standing water, although some species may deposit them on moist substrates near water bodies. The surface location of mosquito eggs serves several crucial functions in the reproductive process. Firstly, positioning the eggs on the water surface provides protection from drowning and predation, as it allows for easy access to atmospheric oxygen for respiration [9]. Additionally, the surface tension of the water supports the buoyancy of the eggs, preventing them from sinking and facilitating their development [47].

The exchange of oxygen is vital for the survival and development of mosquito eggs. Like all insect eggs, mosquito eggs require oxygen for respiration during embryonic development. The thin outer layer of the egg, called the chorion, is permeable to gases, allowing oxygen to diffuse into the egg while carbon dioxide is released [9]. This exchange of gases through the eggshell is essential for maintaining aerobic respiration and ensuring the viability of the developing embryo.

Various household substances have been identified for their efficacy in destroying mosquito eggs, ranging from natural and environmentally friendly options to chemical compounds with larvicidal properties. Soap disrupts water surface tension, suffocating mosquito eggs, while vinegar creates an inhospitable acidic environment hindering egg development. Essential oils like citronella possess larvicidal properties, deterring egg-laying and destroying existing eggs, and diluted bleach solutions disrupt larvae and egg integrity. Additionally, a thin layer of vegetable oil forms a barrier on water surfaces, suffocating eggs. However, effectiveness can vary based on factors like concentration and application method, emphasizing the need for caution, especially with chemical-based products, to ensure safety for humans and non-target organisms [48,36,9].

## 3. MATERIALS AND METHODS

### 3.1 The Human-Controlled Breeding Site (HBCS) method

The HCBS method represents a proactive approach to controlling mosquito populations through targeted intervention at key stages of the mosquito life cycle. This section delineates the step-by-step methodology for implementing the HCBS method, emphasizing its efficacy in attracting gravid female mosquitoes and disrupting the mosquito reproductive process.

#### 3.1.1 Deployment of HCBS Devices

The first step in the HCBS method involves the strategic placement of HCBS devices in areas susceptible to mosquito breeding in urban and suburban environments. HCBS devices are positioned at floor level to maximize visibility to flying female mosquitoes while remaining inconspicuous to human observation. The devices are distributed evenly across the target area to ensure comprehensive coverage and efficacy in mosquito population control.

To minimize breeding site competition effectively, two strategies are essential: (a) the elimination of existing breeding sites, and/or (b) the incorporation of other standing water sources into the HCBS system. In the former approach, the nearest breeding site would be the HCBS device, compelling gravid mosquitoes to utilize it exclusively. Conversely, in the latter approach, regulating potential standing water sources to align with the HCBS methodology transforms them into functional HCBS devices.

#### 3.1.2 Attracting Gravid Female Mosquitoes

Once installed, HCBS devices are designed to attract gravid female mosquitoes seeking oviposition sites for eggs that must be laid. The design features of the HCBS devices, including a shallow basin and a portion of the water surface covered from light, mimic natural breeding habitats favored by mosquitoes. This strategic design encourages female mosquitoes to lay their eggs in the HCBS devices, thereby initiating the mosquito life cycle within a controlled environment.

#### 3.1.3 Removal and Treatment of Water

After a predetermined period, typically 10 days to coincide with the development of mosquito larvae into pupae, the water from the HCBS devices is carefully removed and transferred to a separate container. This step is crucial since if the water is not removed within this timeframe, mosquito pupae will emerge as adults, rendering the method ineffective and inadvertently contributing to mosquito reproduction. Moving forward, it is imperative to explore the feasibility of automating the water replacement process in HCBS devices to mitigate this issue and ensure the continued efficacy of mosquito population control efforts.

#### 3.1.4 Destroying and killing the collected eggs, larvae and pupas

One option to destroy and kill the eggs, larvae and pupas from the HCBS devices, it’s to treat it with household substances known for their larvicidal properties. Common substances such as soap, vinegar, essential oils, or bleach are added to the water to destroy mosquito eggs, larvae, and pupae present in the container. This targeted treatment ensures the eradication of mosquito offspring and prevents their proliferation in the environment (see Section 2.5.5).

It is crucial to emphasize that only water should be poured into the HCBS container. If the mosquito female detects traces of other substances, including insecticides or household products discussed previously, there is a risk that she may reject the HCBS device as an oviposition site. This would render the method ineffective and unsuitable for mosquito population control.

In dire scenarios where household substances are unavailable, simply pouring the water from the HCBS device onto a dry surface can effectively eliminate larvae and pupae but not necessary eggs. However, the easiest alternative to kill the individuals on the three stages is just to bury the collected water beneath a few centimeters of soil (in a garden or even a pot), thus ensuring continued mosquito population control.

#### 3.1.5 Reinstallation of HCBS Devices

Following treatment, the HCBS devices are refilled with fresh water and reinstated in their original location.

### 3.2 Considerations taken during the design of the HCBS Device

The design of the Human-Controlled Breeding Site (HCBS) device was informed by a comprehensive literature review on mosquito ethology and its interactions with humans (Section 2). Understanding the behavior of mosquitoes and their preferences for oviposition sites needs was crucial in developing an effective breeding site for controlling mosquito populations.

#### 3.2.1 Shape Design

The HCBS device was designed with a shallow basin to mimic natural breeding habitats favored by mosquitoes. Its shape allows for a relatively large-exposed water surface, providing ample space for egg deposition. Additionally, the device incorporates a portion of the water surface covered from light, offering protection to mosquito offspring against potential predators.

#### 3.2.2 Color Selection

Five colors were selected for testing the HCBS device: White, Black, Red, Green, and Blue. Color selection played a significant role in determining the attractiveness of the device to gravid female mosquitoes. The color of the HCBS device was chosen to ensure visibility to mosquito females while remaining semi-hidden to humans. This balance is crucial to ensure that mosquitoes perceive the oviposition site as hidden from predators.

#### 3.2.3 Placement and Visibility

The HCBS device is ideally located at floor level to ensure maximum visibility from flying female mosquitoes seeking oviposition sites. It must be strategically positioned to be visible to mosquito females while remaining semi-hidden from human view, creating the perception of a secure oviposition place away from predators.

#### 3.2.4 Additional Features

To facilitate transportation and maintenance, the HCBS device is equipped with a cap or slicing cover to close the exposed water surface when not in use or during harvesting. This feature ensures the integrity of the breeding site and prevents contamination. Importantly, no insecticides or mosquito-killing devices are added to the water within the HCBS to avoid deterring gravid female mosquitoes from ovipositing.

### 3.3 Experiment design

The experiment was conducted in Peru, specifically in the department of Lima, within the district of La Molina, which is situated at an altitude of approximately 237 meters above sea level. Its geographical coordinates are approximately 12.0760° S latitude and 76.9502° W longitude. This region experiences climatic conditions typical of a coastal desert, characterized by moderate temperatures, high relative humidity, and minimal rainfall. During the months of February to May, which coincide with the austral summer, La Molina typically records average temperatures ranging from 22°C to 27°C. Relative humidity levels vary between 60% to 70%. Rainfall is scarce, with an average total annual precipitation of less than 10 millimeters.

The mosquito species commonly found in this area include *Aedes aegypti*, the primary vector for diseases such as dengue, Zika, and chikungunya. Additionally, *Culex quinquefasciatus*, known for transmitting West Nile virus, and *Anopheles* species responsible for malaria transmission, are also prevalent in the region.

The experiment was designed to evaluate the efficacy of Human-Controlled Breeding Site (HCBS) devices in attracting gravid female mosquitoes for oviposition, with a focus on color preference. The HCBS devices were specially designed based on findings from the literature review on mosquito ethology and breeding site preferences. Using 3D printing technology, the devices were fabricated in ABS material in five different colors: white, black, red, green, and blue.

#### 3.3.1 Installation and Setup

All HCBS devices were filled to the middle with potable water and installed in the same location, following the specified placement guidelines to ensure maximum visibility to flying female mosquitoes. After 10 days of exposure to mosquito access, the HCBS devices were capped and removed from their location. To minimize potential biases due to the very specific location of each HCBS device within the installation location, the distribution of colors among HCBS devices was randomly altered at each reinstallation. This process was replicated five times to ensure robustness and reliability of the findings.

#### 3.3.2 Data Collection and Analysis

In the laboratory, a count of larvae and pupae from each HCBS device was conducted to evaluate mosquito attraction. Due to equipment constraints, egg counts were not feasible for this experiment, and therefore, only larvae and pupae were included in the tally. After counting, the HCBS devices were refilled with water and reinstalled in their original location.

It’s worth noting that in our study, we prioritized assessing the functionality of HCBS rather than eradicating the mosquito population in our test area. Consequently, the eggs, larvae, and pupae were not eradicated after collection; instead, they were transferred to another container to mature into adults and contribute to the ongoing mosquito population in the study area. This approach ensured a consistent supply of adult mosquitoes for testing the effectiveness of HCBS methodology and devices.

#### 3.3.3 Laboratory Environment

The installation site was situated inside a laboratory exposed to mosquito access, providing a semi-controlled environment for the experiment (conditions similar to those of a room in a house). This setting allowed for consistent conditions across replicates and minimized external variables that could influence mosquito behavior and oviposition decisions.

## 4. RESULTS AND DISCUSSION

### 4.1 The design

Figure 1 shows the final design of the HCBS device used in our study whereas Figure 2 shows the different color devices installed during the experiment.

**Figure 1:**
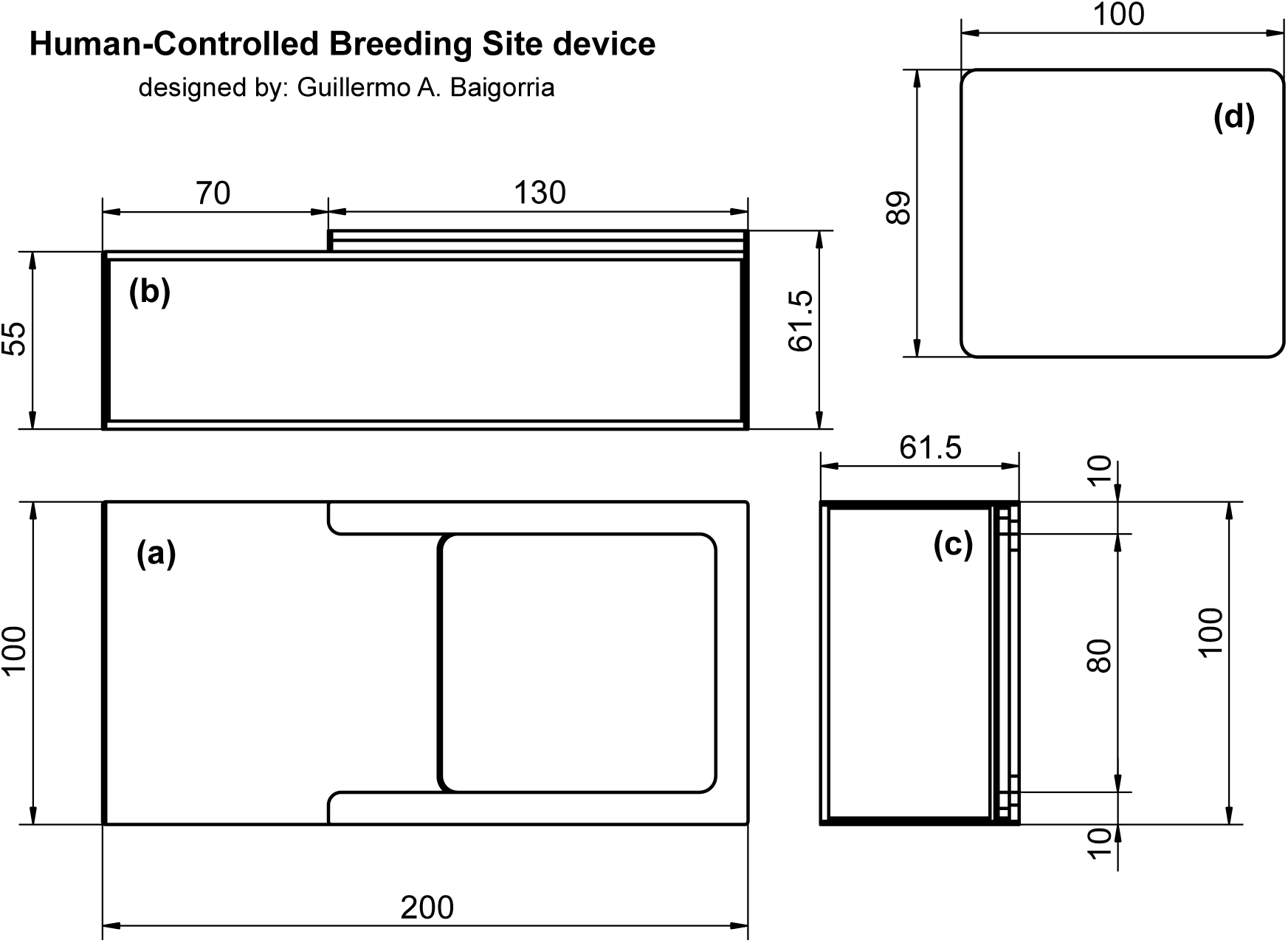
Design drawings of the HCBS device, showing (a) top view of the body, (b) side view of the body, (c) front view of the body, and (d) top view of the slicing cap. Measurements are in millimeters (mm).

**Figure 2:**
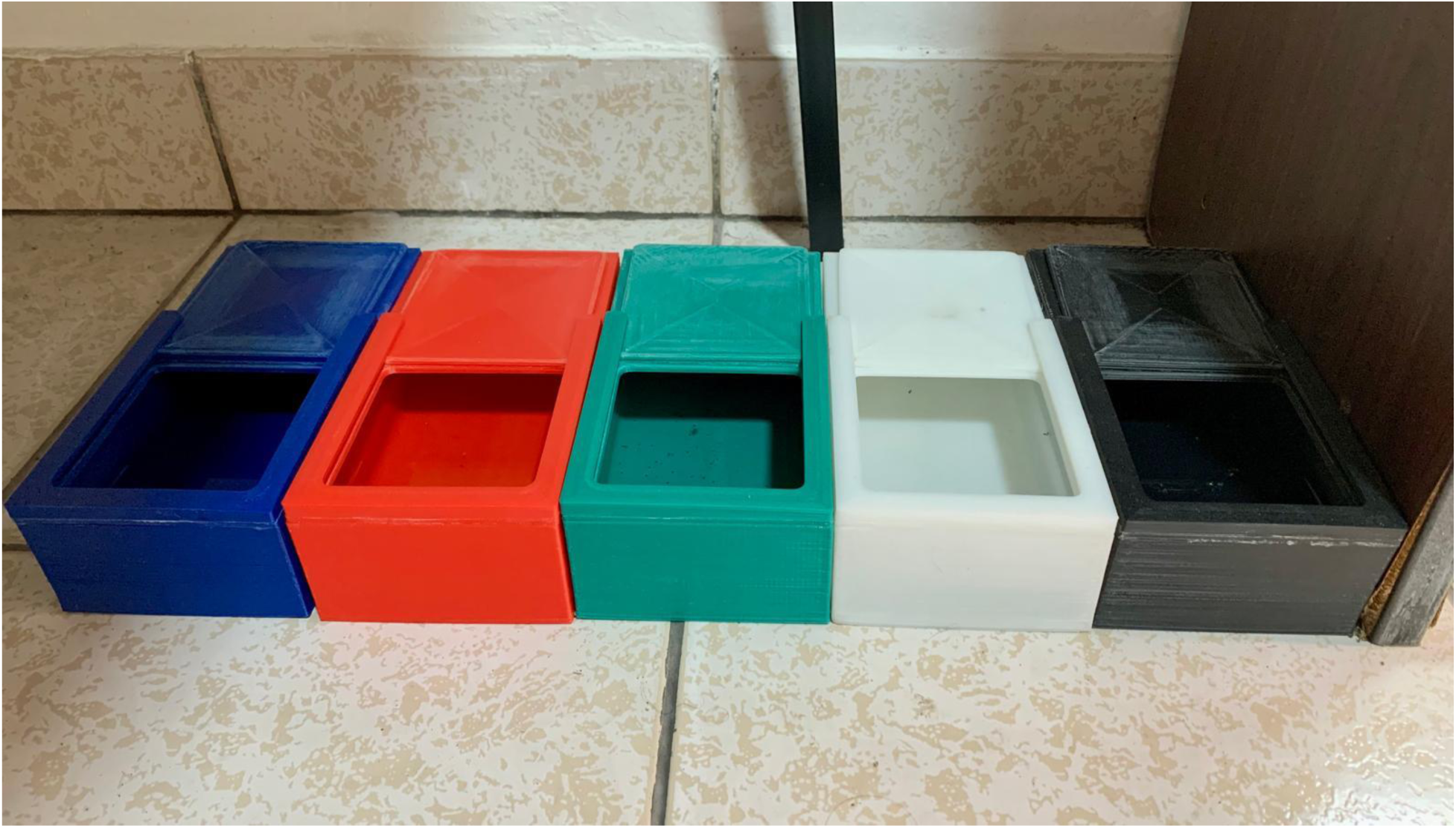
Photograph depicting HCBS devices in all colors used during the experiment, installed in their respective locations.

The slicing cap (d) is placed on the top view of the body (a), enabling slicing between both arms for easy opening and closing of the HCBS device.

### 4.2 Experiment results

#### 4.2.1 Results on the HCBS method

The successful deposition of eggs by gravid mosquitoes into the HCBS devices, followed by the destruction of these individuals within the 10-day timeframe, serves as compelling evidence of the efficacy of the system. This crucial demonstration underscores the functionality of both the HCBS method and device. Without the presence of HCBS devices, these eggs would have inevitably hatched, contributing to the burgeoning mosquito population. Thus, the ability to attract gravid mosquitoes and disrupt their reproductive cycle within the designated timeframe serves as a robust validation of the HCBS system’s effectiveness. This innovative approach challenges the commonly current recommendation to destroy all possible locations where mosquitoes can lay their eggs, providing a controlled environment for oviposition within households while simultaneously reducing mosquito populations at the source.

#### 4.2.2 Results on the HCBS color device

The sampled adult mosquito population from the laboratory revealed that all captured mosquitoes belonged to the *Aedes aegypti* species.

After tallying the larvae and pupae across all HCBS devices, a significant statistical difference was observed in the number of specimens collected, particularly between the green HCBS device and those of other colors. This difference can be attributed to mosquitoes’ exclusive initial preference for ovipositing in the green device, as detailed in Table 1. Counting ceased after 50 days when no oviposition was detected in HCBS devices of other colors, although the devices remained operational in the laboratory. However, subsequent inspections revealed the presence of larvae in the red HCBS device. Two weeks later, larvae were also found in the white device, followed by the black device a week after, and eventually in the blue device. Although the counting had already stopped, preventing precise quantification of individuals, it is crucial to note these later observations. These findings suggest that while green is the preferred color for oviposition, it is not an indispensable condition for gravid mosquitoes to oviposit.

**Table 1:** Number of mosquito’s larvae and pupas in the HCBS devices by color and date:

| HCBS<br>device color | Harvesting date |  |  |  |  |
| --- | --- | --- | --- | --- | --- |
|  | April 20<sup>th</sup> | April 30<sup>th</sup> | May 10<sup>th</sup> | May 20<sup>th</sup> | May 30<sup>th</sup> |
| Black | 0 | 0 | 0 | 0 | 0 |
| Blue | 0 | 0 | 0 | 0 | 0 |
| Green | 2 | 8 | 17 | 15 | 12 |
| Red | 0 | 0 | 0 | 0 | 0 |
| White | 0 | 0 | 0 | 0 | 0 |

The Human-Controlled Breeding Sites (HCBS) method demonstrates remarkable efficacy in controlling mosquito populations, exerting influence over three of the four life stages of mosquitoes. Our findings confirm that when HCBS is correctly applied, it achieves a 100% effectiveness in disrupting the mosquito life cycle by collecting laid egg and eliminating all together with larvae and pupae previously hatched.

One of the key advantages of HCBS is its low implementation cost, which promotes widespread citizen participation in mosquito reduction campaigns. This cost-effectiveness not only encourages community engagement but also facilitates the scalability of mosquito control efforts in urban and suburban areas.

In the event of massive HCBS deployment in urban and suburban environments, a significant reduction in mosquito populations can be anticipated. By disrupting reproduction, HCBS ensures that the current adult mosquito population will be the last local generation. Within a short timeframe, typically within 1 week to 10 days, the absence of male mosquitoes prevents fertilization, effectively limiting the population’s ability to replenish since only unfertilized female mosquitoes would theoretically remain alive after this period.

While the primary focus of HCBS implementation is in urban and suburban areas, its potential applicability in rural settings warrants consideration. Gravid female mosquitoes tend to seek the closest and safest place to lay their eggs. With HCBS devices installed within or around houses, mosquitoes are likely to prefer ovipositing in the HCBS rather than expending energy to find distant natural environments where predators exist (see Section 2.5.5). Although its effectiveness may be somewhat diminished compared to urban areas, HCBS still serves to reduce local mosquito populations and disrupt the feedback loop of mosquito repopulation in natural environments.

Our study also identifies the importance of device color selection in optimizing HCBS effectiveness. Among the colors tested, green HCBS devices consistently demonstrated the highest efficacy in attracting gravid female mosquitoes for oviposition; however, it is not an indispensable condition. Additionally, HCBS devices must have a portion of the water surface shielded from light. This feature makes female mosquitoes more likely to select this device for oviposition, as it offers protection for their offspring against predators. This issue was verified during the retrieval and counting of the individuals collected in the laboratory. When the slicing cap of the HCBS device was opened under the lights, all larvae swiftly moved towards the shaded water within the device. This finding underscores the importance of considering mosquito behavior and preferences in the design of effective mosquito control strategies.

Overall, the findings of this study underscore the potential of HCBS as a valuable addition to Integrated Mosquito Management strategies, particularly in urban and suburban settings. By targeting multiple life stages of mosquitoes and incorporating features that enhance attractiveness and efficacy, HCBS offers a promising approach to reducing mosquito populations and mitigating the transmission of mosquito-borne diseases in diverse environments. Further research and field trials are warranted to explore the long-term effectiveness and scalability of HCBS implementation in different geographic contexts. Additionally, optimizing the design of HCBS devices warrants attention to maximize their efficacy and applicability. Finally, the HCBS method can undergo evaluation for potential application in combating other insect-borne vector diseases.

One suggestion is to leverage disaster risk management systems capable of widespread communication via SMS text messages to convey crucial information to residents in a specific area. The proposal involves utilizing a country’s system to coordinate the simultaneous installation of HCBS devices across the entire community or city. Similarly, on the 10^th^ day, residents would receive alerts prompting them to harvest and reinstall the HCBS devices. This approach ensures uniform implementation of control programs throughout the community, reminding residents simultaneously to remove the devices to mitigate the negative effects discussed in Section 3.3.3.

## 5. CONCLUSIONS

The Human-Controlled Breeding Site (HCBS) methodology, developed in Peru but applicable globally, provides a sustainable, low-cost approach to mosquito control by deploying controlled oviposition sites within households to attract gravid female mosquitoes, whose eggs, larvae, and pupae are mechanically destroyed, disrupting three out of the four mosquito life cycle stages. This adaptable method, using household items like cups or bottles, effectively curbs mosquito reproduction in urban and suburban areas, complementing Integrated Mosquito Management and addressing challenges like temephos resistance in Aedes aegypti populations in Peru, where coastal regions show resistance ratios exceeding 20 [49]. By preventing adult emergence within a 10-day cycle, HCBS achieves 100% efficacy, with accessible disposal methods like burying water in soil ensuring applicability even without household substances. Its scalability, supported by community engagement and potential integration with disaster risk management systems, makes HCBS a promising tool to mitigate a wide range of mosquito-borne diseases, including dengue, Zika, West Nile virus, Yellow fever, Chikungunya, and Malaria, enhancing public health outcomes worldwide.

## Data Availability

All data produced in the present work are contained in the manuscript

## Acknowledgements

The authors acknowledge the use of generative AI technology ChatGPT 3.5. to improve the paper’s reading.

## Availability of data and materials

All data is shown on the table and figures presented in the paper.

## Ethics approval and consent to participate

Not applicable

## Consent for publication

Not applicable

## Disclaimer

This paper on Human-Controlled Breeding Sites (HCBS) for mosquito control is intended for research and informational purposes only. The findings, methods, and recommendations presented are based on the authors’ interpretation of available data and scientific literature. While the authors have made every effort to ensure the accuracy and relevance of the content, the information provided does not constitute professional or medical advice. The application of HCBS methods should be conducted under the guidance of qualified experts and in compliance with local regulations and ethical standards. The authors disclaim any liability for the misuse or misapplication of the information contained in this paper, or for any adverse effects resulting from the implementation of the methods discussed. Readers are encouraged to consult with relevant authorities and professionals before undertaking any mosquito control initiatives.

## REFERENCES

1. Mora C, McKenzie T, Gaw IM, Dean JM, von Hammerstein H, Knudson TA, Setter RO, Smith CZ, Webster KM, Patz JA, Franklin EC. Over half of known human pathogenic diseases can be aggravated by climate change. Nat Clim Chang. 2022;12(9):869–875. 10.1038/s41558-022-01426-1.

2. Lafferty KD. The ecology of climate change and infectious diseases. Ecol. 2009;90(4):888–900. 10.1890/08-0079.1

3. Joshi A, Miller C. Review of machine learning techniques for mosquito control in urban environments. Ecol Informatics. 2021;61:101241. 10.1016/j.ecoinf.2021.101241.

4. Guagliardo SA, Barboza JL, Morrison AC, Astete H, Vazquez-Prokopec G, Kitron U. Patterns of geographic expansion of *Aedes aegypti* in the Peruvian Amazon. PLoS Negl Trop Dis. 2014;8(8):e3033. https://doi,org/10.1371/journal.pntd.0003033.

5. Bazin M, Williams CR. Mosquito traps for urban surveillance: collection efficacy and potential for use by citizen scientists. J Vector Ecol. 2018;43(1):98–103. 10.1111/jvec.12288.

6. Rose RI. Pesticides and public health: integrated methods of mosquito management. Emerg Infect Dis. 2001;7(1):17–23. 10.3201/eid0701.010103.

7. Naranjo DP, Qualls WA, Jurado H, Perez JC, Xue R, Gomez E, Beier JC, Vector control programs in Saint Johns County, Florida and Guayas, Ecuador: successes and barriers to integrated vector management. BMC Public Health 2014;14, 674. 10.1186/1471-2458-14-674

8. World Health Organization (WHO). Vector-borne diseases. WHO; 2020 Accessed August 28, 2024. https://www.who.int/news-room/fact-sheets/detail/vector-borne-diseases

9. Clements AN. The Biology of Mosquitoes: Sensory Reception and Behaviour (Vol. 2). CABI; 1999.

10. Lambert B, North A, Charles H, Godfray J. A meta-analysis of longevity estimates of mosquito vector of disease. BioRxiv. 2022;05.30.494059. 10.1101/2022.05.30.494059.

11. Foster WA, Walker ED. Mosquitoes (Culicidae). *In*: Mullen G, Durden L. (eds) Medical and Veterinary Entomology (3^rd^ Edition). Academic Press; 2009. Pages 261–325. 10.1016/B978-0-12-814043-7.00015-7

12. Service MW. Medical Entomology for Students. Cambridge University Press; 2012.

13. Scott TW, Takken W. Feeding strategies of anthropophilic mosquitoes result in increased risk of pathogen transmission. Trends Parasitol. 2012;28(3):114–121. 10.1016/j.pt.2012.01.001.

14. Foster WA, Takken W. Nectar-related vs. human-related volatiles: behavioural response and choice by female and male *Anopheles gambiae* (Diptera: Culicidae) between emergence and first feeding. Bull Entomol Res. 2004;94(2):145–157. 10.1079/ber2003288.

15. Hugo LE, Jeffery JAL, Trewin BJ, Wockner LF, Yen NT, Le NH, Nghia LT, Hine E, Ryan PA, Kay BH, Adult survivorship of the dengue mosquito Aedes aegypti varies seasonally in central Vietnam. PLOS Negl Trop Dis. 2014;8(2):e2669. 10.1371/journal.pntd.0002669.

16. Paaijmans KP, Blanford S, Bell AS, Blanford JI, Read AF, Thomas MB. Influence of climate on malaria transmission depends on daily temperature variation. Proc Natl Acad Sci USA. 2010;107(34):15135–15139. 10.1073/pnas.1006422107.

17. Durrance-Bagale A, Hoe N, Lai J, Liew JWK, Clapham H, Howard N. Dengue vector control in high-income, city settings: A scoping review of approaches and methods. PLoS Negl Trop Dis. 2024;18(4):e0012081. 10.1371/journal.pntd.0012081

18. Gubler DJ. Dengue, urbanization and globalization: the unholy trinity of the 21(st) century. Trop Med Health. 2011;39(4 Suppl):3–11. 10.2149/tmh.2011-S05.

19. Leisnham PT, LaDeau SL, Juliano SA. Spatial and temporal habitat segregation of mosquitoes in urban Florida. PLoS ONE. 2014;9(3):e91655. 10.1371/journal.pone.0091655.

20. Sumba LA, Ogbunugafor CB, Deng AL, Hassanali A. Regulation of Oviposition in *Anopheles gambiae s.s.*: Role of Inter- and Intra-Specific Signals. J Chem Ecol. 2008;34:1430– 1436. 10.1007/s10886-008-9549-5.

21. Allan SA, Kline DL. Larval rearing water and pre-existing eggs influence oviposition by *Aedes aegypti* and *Ae. albopictus* (Diptera: Culicidae). J Med Entomol. 1998;35:943–947. 10.1093/jmedent/35.6.943.

22. Gimnig JE, Ombok M, Otieno S, Kaufman M, Vulule JM, Walker ED. Density-dependent development of *Anopheles gambiae* (Diptera: Culicidae) larvae in artificial habitats. J. Med. Entomol. 2002;39:162–172. 10.1603/0022-2585-39.1.162.

23. Kiflawi M, Blaustein L, Mangel M. Oviposition habitat selection by the mosquito *Culiseta longiareolata* in response to risk of predation and conspecific larval density. Ecol. Entomol. 2003;28:168–173. 10.1046/j.1365-2311.2003.00505.x

24. Koenraadt CJM, Takken W. Cannibalism and predation among larvae of *Anopheles gambiae* complex. Med. Vet. Entomol. 2003;17:61–66. 10.1046/j.1365-2915.2003.00409.x.

25. Allan SA, Kline DL, Larval Rearing Water and Preexisting Eggs Influence Oviposition by *Aedes aegypti* and *Ae. albopictus* (Diptera: Culicidae), J Med Entomol. 1998;35(6): 943–947. 10.1093/jmedent/35.6.943

26. World Health Organization (WHO), the Special Programme for Research and Training in Tropical Diseases (TDR). Dengue guidelines, for diagnosis, treatment, prevention and control. 3^rd^ ed. WHO Press; 2009.

27. Hemingway J, Ranson H. Insecticide resistance in insect vectors of human disease. Annu Rev Entomol. 2000;45(1):371–391. 10.1146/annurev.ento.45.1.371.

28. Sota T, Mogi M. Interspecific variation in desiccation survival time of *Aedes* (*Stegomyia*) mosquito eggs is correlated with habitat and egg size. Oecologia. 1992;90:353–358. 10.1007/BF00317691.

29. Alphey L, Beard CB, Billingsley P, et al. Malaria control with genetically manipulated insect vectors. Science. 2002;298(5591):119–121. 10.1126/science.1078278.

30. Basáñez MG, Razali K, Renz A, Kelly D. Density-dependent host choice by disease vectors: epidemiological implications of the ideal free distribution. Trans R Soc Trop Med Hyg. 2007;101(3):256–69. 10.1016/j.trstmh.2006.08.009.

31. World Health Organization (WHO). Handbook for Integrated Vector Management. WHO Press; 2019.

32. Foster WA, Hancock RG. Nectar-related olfactory and visual attractants for mosquitoes. J Am Mosq Control Assoc. 1994;10(3):288–96.

33. Takken W, Knols BG. Odor-mediated behavior of Afrotropical malaria mosquitoes. Annu Rev Entomol. 1999;44(1):131–157. 10.1146/annurev.ento.44.1.131.

34. Logan JG, Birkett MA, Semiochemicals for biting fly control: Their identification and exploitation. Pest Manag Sci. 2007;63(7):647–57. 10.1002/ps.1408. PMID: 17549674.

35. Alpern JD, Dunlop SJ, Dolan BJ, Stauffer WM, Boulware DR. Personal Protection Measures Against Mosquitoes, Ticks, and Other Arthropods. Med Clin North Am. 2016;100(2):303–16. 10.1016/j.mcna.2015.08.019.

36. Maia MF, Moore SJ. Plant-based insect repellents: a review of their efficacy, development and testing. Malaria J. 2011;10(1):S11. 10.1186/1475-2875-10-S1-S11.

37. Lambrechts L. Scott TW, Gubler DJ. Consequences of the expanding global distribution of *Aedes albopictus* for dengue virus transmission. PLoS Negl Trop Dis. 2010;4(5):e646. 10.1371/journal.pntd.0000646.

38. Sinka ME, Bangs MJ, Manguin S, Coetzee M, Mbogo CM, Hemingway J, Patil AP, Temperley WH, Gething PW, Kabaria CW, Okara RM, Van Boeckel T, Godfray HC, Harbach RE, Hay SI. The dominant *Anopheles* vectors of human malaria in Africa, Europe and the Middle East: occurrence data, distribution maps and bionomic précis. Parasit Vectors. 2010;3(3):117. 10.1186/1756-3305-3-117.

39. Weiss DJ, Lucas TCD, Nguyen M, et al. Mapping the global prevalence, incidence, and mortality of *Plasmodium falciparum*, 2000-17: a spatial and temporal modelling study. Lancet. 2019;394(10195):322–331. 10.1016/S0140-6736(19)31097-9.

40. Bentley MD, Day JF. Chemical ecology and behavioral aspects of mosquito oviposition. Annu Rev Entomol. 1989;34(1):401–421. 10.1146/annurev.en.34.010189.002153.

41. Coutinho-Abreu IV, Riffell JA, Akbari OS. Human attractive cues and mosquito host-seeking behavior. Trends Parasitol. 2021;38(3):246–264. 10.1016/j.pt.2021.09.012.

42. Wong J, Stoddard ST, Astete H, Morrison AC, Scott TW. Oviposition site selection by the dengue vector Aedes aegypti and its implications for dengue control. PLoS Negl Trop Dis. 2011;5(4):e1015. 10.1371/journal.pntd.0001015.

43. Munga S, Minakawa N, Zhou G, Barrack OO, Githeko AK, Yan G. Oviposition site preference and egg hatchability of Anopheles gambiae: effects of land cover types. J Med Entomol. 2005;42(6):993–7. 10.1093/jmedent/42.6.993.

44. Lounibos LP. Invasions by insect vectors of human disease. Annu Rev Entomol. 2002;47(1):233–66. 10.1146/annurev.ento.47.091201.145206.

45. Service MW. Mosquito Ecology: Field Sampling Methods. Elsevier; 1997.

46. Christophers, S.R. (1960) Aedes aegypti (L.), the Yellow Fever Mosquito: Its Life History, Bionomics and Structure. Cambridge University Press, London, 739 p.

47. Clements AN. The Biology of Mosquitoes: Development, Nutrition and Reproduction (Vol. 1). CABI; 2000.

48. Fernando HSD, Saavedra-Rodriguez K, Perera R, Black WC 4th, De Silva BGDNK. Resistance to commonly used insecticides and underlying mechanisms of resistance in Aedes aegypti (L.) from Sri Lanka. Parasit Vectors. 2020;13(1):407. 10.1186/s13071-020-04284-y.

49. Palomino M, Pinto J, Yañez P, Cornelio A, Dias L, Amorim Q, Queiroz A, Serrano JC, Lenhart A, Conn JE. First national-scale evaluation of temephos resistance in *Aedes aegypti* in Peru. Parasit Vectors. 2022;15(1):254. 10.1186/s13071-022-05310-x

